# Going by the Numbers : Learning and Modeling COVID-19 Disease Dynamics

**DOI:** 10.1101/2020.05.18.20106112

**Authors:** Sayantani Basu, Roy H. Campbell

## Abstract

The COrona VIrus Disease (COVID-19) pandemic caused by the severe acute respiratory syndrome coronavirus 2 (SARS-CoV2) has resulted in a challenging number of infections and deaths worldwide. In order to combat the pandemic, several countries worldwide enforced mitigation measures in the forms of lockdowns, social distancing and disinfection measures. In an effort to understand the dynamics of this disease, we propose a Long Short Term Memory (LSTM) based model. We train our model on nearly four months of cumulative COVID-19 cases and deaths. Our model can be adjusted based on the parameters in order to provide predictions as needed. We provide results at both the country and county levels. We also perform a quantitative comparison of mitigation measures in various counties in the United States based on the rate of difference of a short and long window parameter of the proposed LSTM model. The analyses provided by our model can provide valuable insights based on the trends in the rate of infections and deaths. This can also be of help for countries and counties deciding on mitigation and reopening strategies. We believe that the results obtained from the proposed method will contribute to societal benefits for a current global concern.

## 1 Introduction

The outbreak of a novel coronavirus which spread from China in December 2019 led to severe health problems and fatalities worldwide, eventually turning into a global pandemic. The virus was named the severe acute respiratory syndrome coronavirus 2 (SARS-CoV2) and the disease caused due to it was named the COrona VIrus Disease (COVID-19). The most common symptoms are cough and fever, although symptoms range in severity including pneumonia-like symptoms and dyspnea [1]. It was confirmed that COVID-19 spread through infected respiratory droplets transmitted from an infected person to a healthy person [2]. Other symptoms in humans have been studied, including relations to the gastrointestinal system [3] and skin rashes [4]. More recent studies have focused on other possible zoonotic pathways, including other species that can possibly contract COVID-19 [5].

Till date, there is no definite medical treatment for COVID-19, although global efforts are in progress for development of a vaccine or specific drugs. Current methods for treatment have involved the use of antiviral medications commonly used for viral diseases [6, 7, 8].

Healthcare professionals were given special instructions and equipment in order to treat patients who tested positive for COVID-19. There was also a surge in demand for PPE (Personal Protective Equipment), especially for frontline healthcare workers as well as for ventilators in hospitals [9, 10].

Countries all over the world executed various mitigation measures for the COVID-19 pandemic, including social distancing, enforcing total lockdowns, and advising people to take precautions like wearing face masks and washing hands. These measures were implemented based on projection studies and statistical models that showed the effect of mitigation measures [11, 12, 13].

Usually individuals showing common COVID-19 symptoms were asked to self-isolate for two weeks and seek medical help if health conditions worsened [14, 15, 16]. COVID-19 was generally observed to affect specific groups of people in populations [17, 18], as well as people who have pre-existing medical conditions [19, 20].

It was noted that people may be capable of transmitting the virus despite being completely asymptomatic or in other words, testing positive for COVID-19 without any visible symptoms [21]. This may have expedited the spread of the disease [22] as certain individuals may have been unaware of their conditions and symptoms and transmitted the disease while socializing before they medically tested positive for COVID.

In the present work, we propose an LSTM based approach to understand and evaluate the effectiveness of mitigation measures at both the country and county levels. The rest of this paper is organized as follows: Section 2 discusses the related work, Section 3 discusses the proposed method, Section 4 discusses the results, and Section 5 concludes the paper and suggests future work.

## 2 Related Work

The spread of COVID-19 led to global research efforts to understand the disease and its possible implications. We focus on discussing related work with regard to COVID-19 disease dynamics, especially studies involving simulations and computational learning algorithms.

Several studies have focused on projections of how the infections are likely to spread based on statistical analyses. Neher et al. [23] have proposed a seasonal transmissability model based on SIR [24], a class of mathematical epidemiological models to predict the trajectory of the virus re-infecting sub-populations in the world in future years. They have taken into account the infection, emigration and population turnover rates respectively as part of their proposed model. A refined version of their model [25] considers visualizing the projected hospital resources necessary in order to support a pandemic like COVID-19.

Another similar hospital impact model CHIME (COVID-19 Hospital Impact Model for Epidemics) developed by Penn Medicine [26] suggests the projection of the hospitalized, ICU (Intensive Care Unit) and ventilated patients respectively for Penn hospitals. They have used a statistical model based on measures like social contact, hospitalization rate and probability of detection.

With regard to COVID-19, there have been several machine learning based models proposed by various research teams globally in an effort to understand the different facets of COVID-19.

Hu et al. [27] proposed a machine learning model based on stacked auto-encoders to forecast cumulative COVID-19 cases in China till April 2020 based on past cumulative data. They also grouped the cities and provinces in clusters based on the features extracted from their proposed auto-encoder model. Similar LSTM approaches have also been explored in order to understand patient related data [28] as well as to study epidemic trends in China [29].

All COVID-19 forecast and projection models proposed so far have been more focused on graphical analyses and future curve prediction. However, in our proposed model we take quantitatively provide an evaluation of the degree to which mitigation measures are working based on the future epidemiological curves for COVID-19.

Wang et al. [30] have proposed an “Ontology-based Side-effect Prediction Framework (OSPF)” for evaluating the “Traditional Chinese Medicine (TCM)” model using deep learning. They have trained an Artificial Neural Network (ANN) model with an architecture composed of three hidden layers on the TCM prescriptions used for the treatment of COVID-19 patients. They have also tested the validity of their OSPF model to evaluate the effectiveness and safety of use of TCM prescriptions for other flu-like diseases as a potential source of treatments for COVID-19. Their proposed model classifies a given TCM prescription as ‘Safe’ or ‘Unsafe’. They have identified seven TCM prescriptions that should take precedence over others while treating COVID-19 patients.

Wang et al. [31] have proposed BI-AT-GRU, a GRU (Gated Recurrent Unit) based model with bidirectionality and attention in order to classify six different respiratory patterns. They have specifically observed that abnormal breathing patterns in patients diagnosed with COVID-19 arise significantly due to Tachypnea, which is characterised as abnormally rapid and shallow breathing. They show that their proposed model is able to distinguish respiratory patterns of ‘Tachypnea’, ‘Central-Apnea’, ‘Bradypnea’, ‘Eupnea’, ‘Cheyne-Stokes’ and ‘Biots’ with high F-1 scores.

Ying et al. [32] have proposed a deep learning model for studying CT (Computer Tomography) images of COVID-19 patients. They collected lung CT images from patients having COVID-19, bacterial pneumonia and healthy people respectively from two hospitals in China. Their proposed framework DeepPneumonia has an AUC (Area Under Curve) metric of 0.99 and sensitivity of 0.93 with regard to distinction of COVID-19 from other diseases.

Wang et al. [33] have proposed a CNN (Convolutional Neural Network) based model ‘Inception Migration Neuro Network’ to identify the radiographical features in volumetric chest CT images of patients obtained from two medical institutions in China. The aim of their proposed model is to identify whether COVID-19 is present or not in the CT images of the patients. They have evaluated AUC and F-1 metrics on their model and have claimed that their method is not invasive and is low cost.

Li et al. [34] have proposed a deep learning framework ‘COVID-19 detection neural network’ or ‘COVNet’ to distinguish CT scans of COVID-19 patients from those of ‘CAP (Community Acquired Pneumonia)’ and other non-pneumonia diseases. They have proposed a three-dimensional model involving ResNet50, max-pooling and a fully connected layer to identify the presence of COVID-19 in the images.

Sethy and Behera [35] have proposed a similar machine learning model for X-Ray images based on ResNet50 and SVM (Support Vector Machines) in order to identify patients with COVID-19. They have evaluated the efficiency of their proposed model using False Positive Rate (FPR), F1 score, Kappa and MCC (Matthews correlation coefficient) metrics.

In the present work, we explore the possibilities of using an LSTM-based approach to learn patterns in COVID-19 data. Statistical learning models require thorough understanding of data and coefficient tuning in order to fit models suitable for prediction. However, as opposed to statistical learning, machine learning approaches involve allowing the model to automatically learn complex patterns from the data based on the model constructed and tuned hyperparameters. This is particularly useful for handling epidemiological time-series data like that of COVID-19 where the cumulative curves of infections and deaths vary with respect to place and time.

## 3 Proposed Method

Long Short-Term Memory Networks (LSTMs) that were originally proposed by Hochreiter and Schmidhuber [36], are a special kind of Recurrent Neural Networks (RNNs) that have the ability of capturing long-term dependencies. In this paper, we apply LSTMs to study the disease dynamics of COVID-19, specifically in forecasting expected numbers of cases, deaths and recoveries. Most importantly, the capabilities of LSTMs allow us to gain a deeper understanding of where the model stands in terms of future predictions and will serve as an aid in taking important healthcare and mitigation measures.

### 3.1 Dataset

For the purpose of training, validation and testing, we utilize data from the Johns Hopkins University repository [37]. For experimental purposes, we consider data over the period from January 22 to June 10. The dataset provides time-series data of COVID-19 confirmed cases, deaths and recovered cases. We separately consider each of these scenarios, as well as look into more fine-grained data at the county-level in order to analyse the effectiveness of mitigation measures.

Separating time series data into train, validation and test sets requires care since all the temporal relations have to be maintained. In other words, it would be unfair to let the model randomly predict the number of confirmed cases at time *t* + 1 if it has not been trained on the observation at time *t* and possibly other time steps before that. At the same time, it is essential to monitor the generalization of and select hyperparameters for the model accordingly. In order to handle this, we partition the data based on time for training and validation as shown in Figure 1.

**Figure 1:**
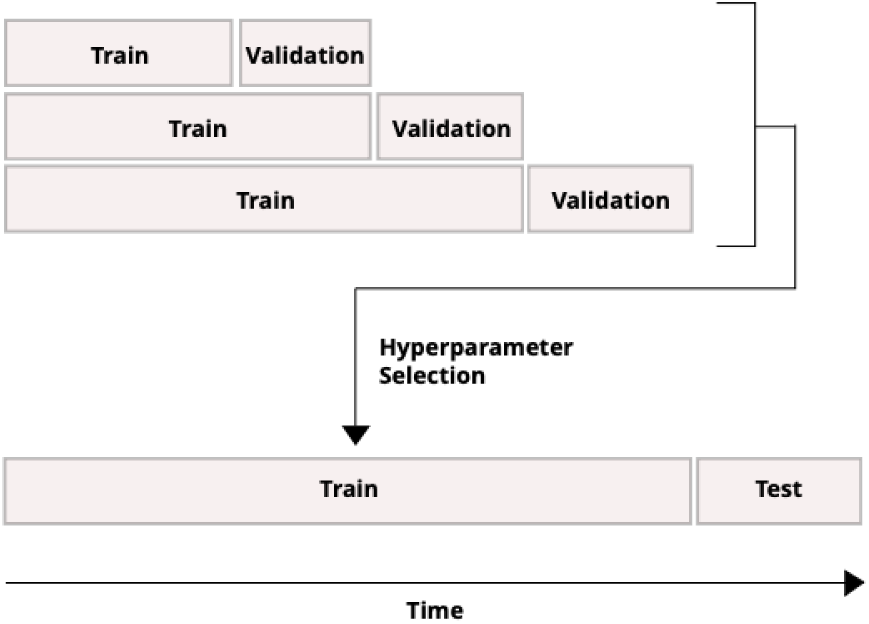
Dataset split for LSTM model.

The data is separately trained on three different train and validation splits and hyperparameters with minimum validation root mean squared error (RMSE). The hyperparameters we specifically focus on tuning are the *lookback* and *lookahead*. The split of train and test data for the tuned model is approximately maintained at an 80:20 ratio based on the initial *lookback* and *lookahead* of 1 and 1 respectively, although the model is capable of handling other splits based on the data. The smaller splits of train and validation sets are approximately maintained at a 60:20 ratio. Intuitively, an increase in the *lookback* and *lookahead* parameters reduces the number of training samples and testing samples respectively and leads to erroneous results. For experimentation purposes, we test *lookback* in the range of 1 and 7 and *lookahead* in the range of 5 and 7 for reasonable future predictions, while adhering to the index constraints on individual train, validation and test data.

### 3.2 Model Design

For the proposed LSTM model, Keras was used as the framework with TensorFlow as backend.

In all our experiments, we train the LSTM for 10,000 epochs with 10 layers and a batch size of 10. All normalization is performed on the *log* scale and Adam [38] is used as the optimizer with a learning rate of 0.001. The window parameters we specifically focus on in this case are the *lookback* and *lookahead* parameters which are tuned based on the data of a particular location. In this case, the *lookback* parameter is indicative of the window size in terms of the number of consecutive days the LSTM needs to “look back” at in order to make a reasonable prediction. The *lookahead* parameter indicates the number of days in advance that the system needs to “look ahead” in order to make a prediction for a future date.

In general, the model is able to provide future predictions of confirmed cases/deaths. However, from a research perspective, we plot the predictions obtained after training on the suitable hyperparameters by comparing them with the actual values of confirmed cases/deaths observed on those specific days for obtaining useful interpretations. The detailed analyses of the results and corresponding figures are discussed in Section 4.

## 4 Results and Discussion

Our analyses based on our proposed LSTM model are divided into two main parts: the prediction models that show predictions of COVID-19 confirmed cases/deaths and evaluations on how well social distancing/mitigation measures are working in the present scenario. In all our plots, dates in January, February, March, April, May, and June are prefixed with ‘J’, ‘F’, ‘M’, ‘A’, ‘MA’, and ‘JU’ respectively followed by the day of the month. As aforementioned, all modeling is carried out based on the data provided for a specific location.

### 4.1 Prediction Models

We discuss our results on two levels: country-level and county-level data on COVID-19 confirmed cases and deaths. In addition to plotting the predicted values and actual values, we also plot a baseline in all figures that is calculated as the moving average over the actual values in the *lookback* window. Overall, it was observed that it was harder to predict the number of comfirmed cases compared to the number of deaths. It was also observed in general that shorter values of *lookback* and *lookahead* resulted in lower values of both the train and test RMSE. However, in the present situation, it is of more interest to discuss the interpretation of what the LSTM predictions are indicative of instead of purely focusing on the values of the RMSE.

#### 4.1.1 Country-level Predictions

Figure 2 shows the plot of predictions for COVID-19 confirmed cases in the United States of America. The United States had different lockdown and phased reopening strategies implemented in different parts of the country [39]. The LSTM predictions for COVID-19 cases suggest that the cumulative cases are still increasing. However, the actual number of COVID-19 cases in the entire nation also seems to be increasing despite some states implementing social distancing measures as early as March. This suggests that mitigation measures are working but the curve for the entire nation is yet to be flattened.

**Figure 2:**
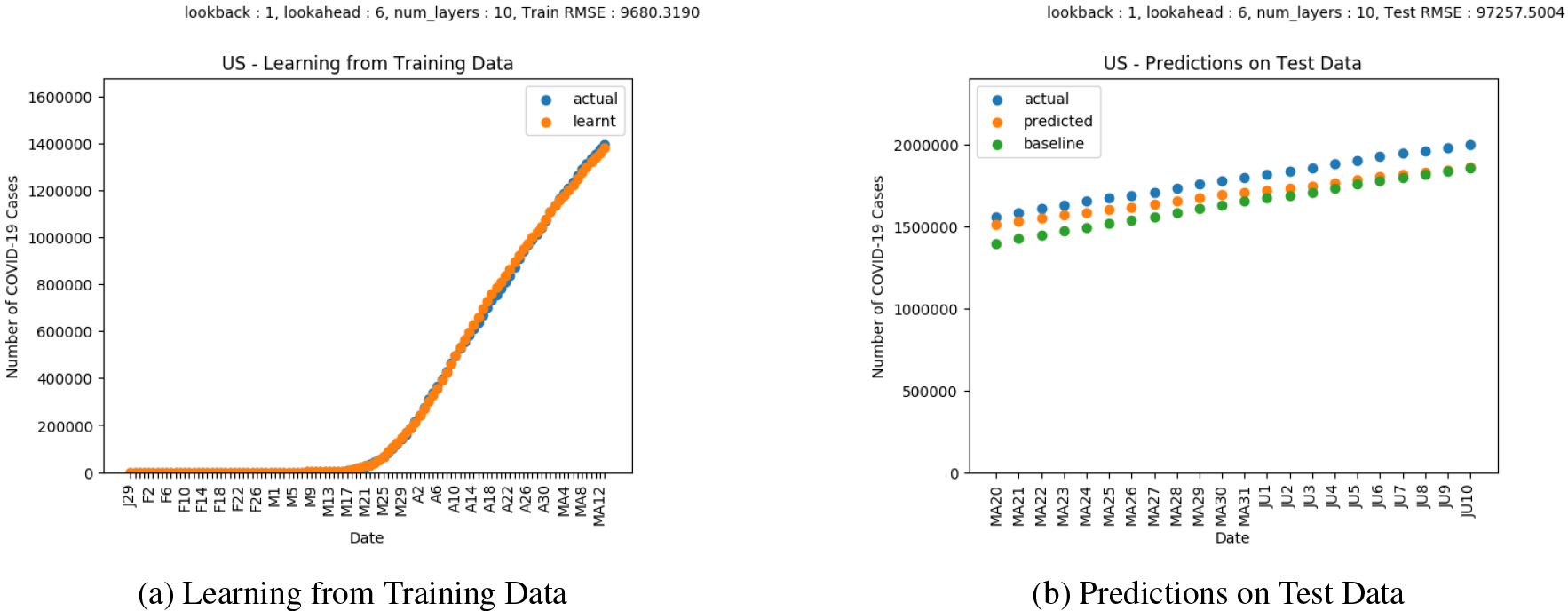
Results on COVID-19 Cases in the United States of America.

Figure 3 shows the plot of predictions for COVID-19 deaths in United States of America. The LSTM predictions in this scenario show a trend similar to the cumulative COVID-19 cases. However, the predictions suggest that the deaths would continue to increase but at a lower rate.

**Figure 3:**
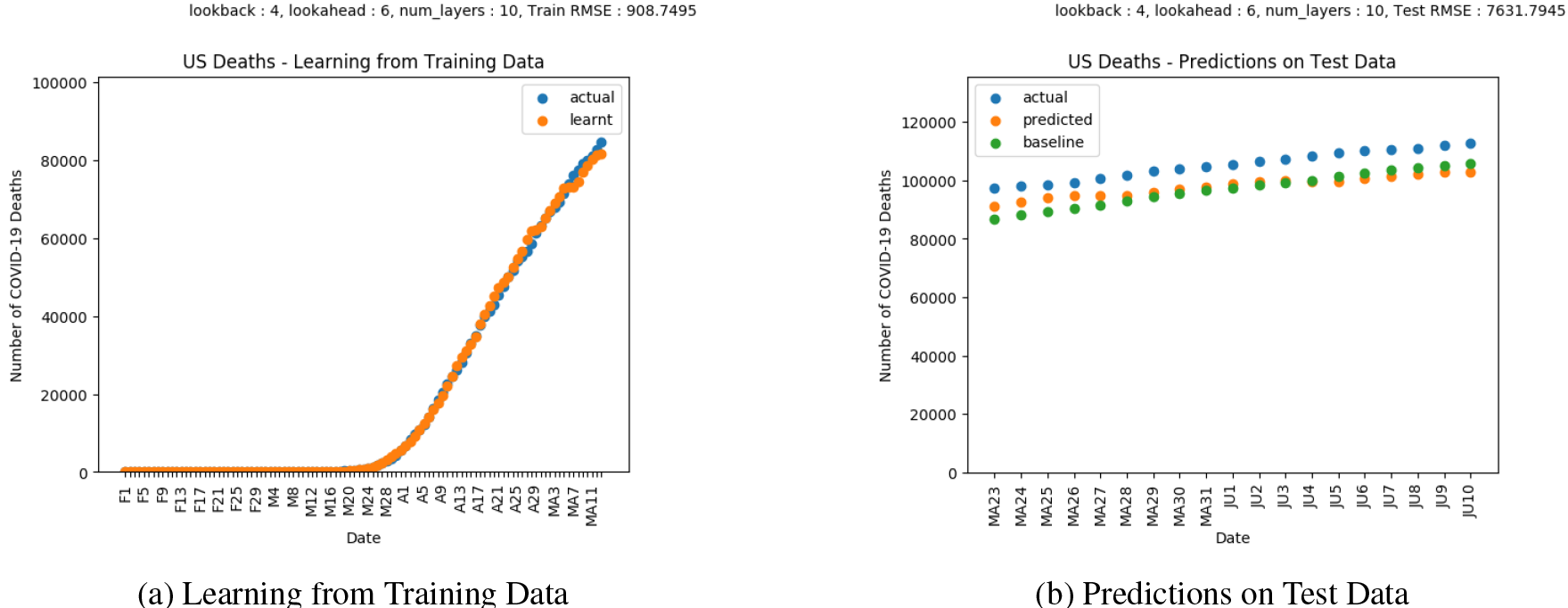
Results on COVID-19 Deaths in the United States of America.

Figure 4 shows the plot of predictions for COVID-19 confirmed cases in Italy. Italy progressively enforced strict lockdown policies in various regions starting February 23 [40]. The LSTM predictions follow a similar trend with regard to the actual values of COVID-19 cases, and is able to forecast the flattening of the curve achieved by Italy.

**Figure 4:**
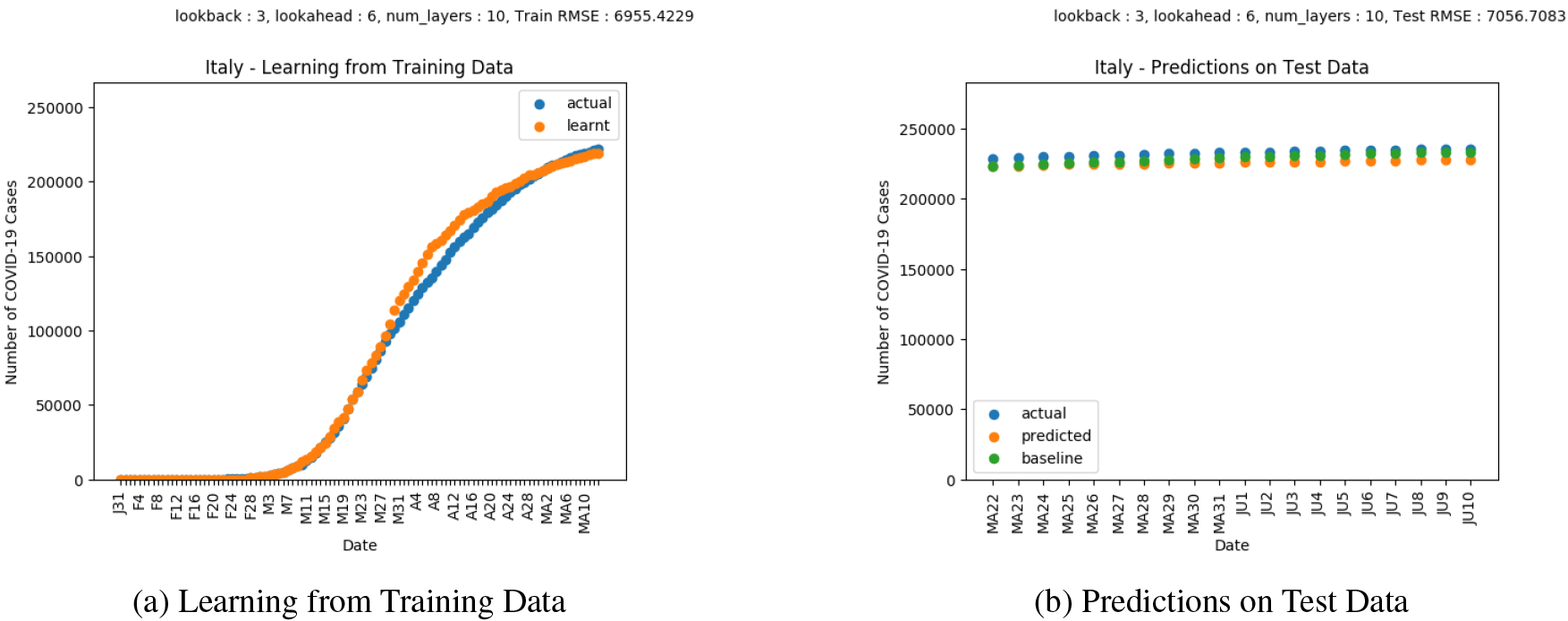
Results on COVID-19 Cases in Italy.

Figure 5 shows the plot of predictions for COVID-19 deaths in Italy. The deaths in Italy show a similar trend as that of the cases.

**Figure 5:**
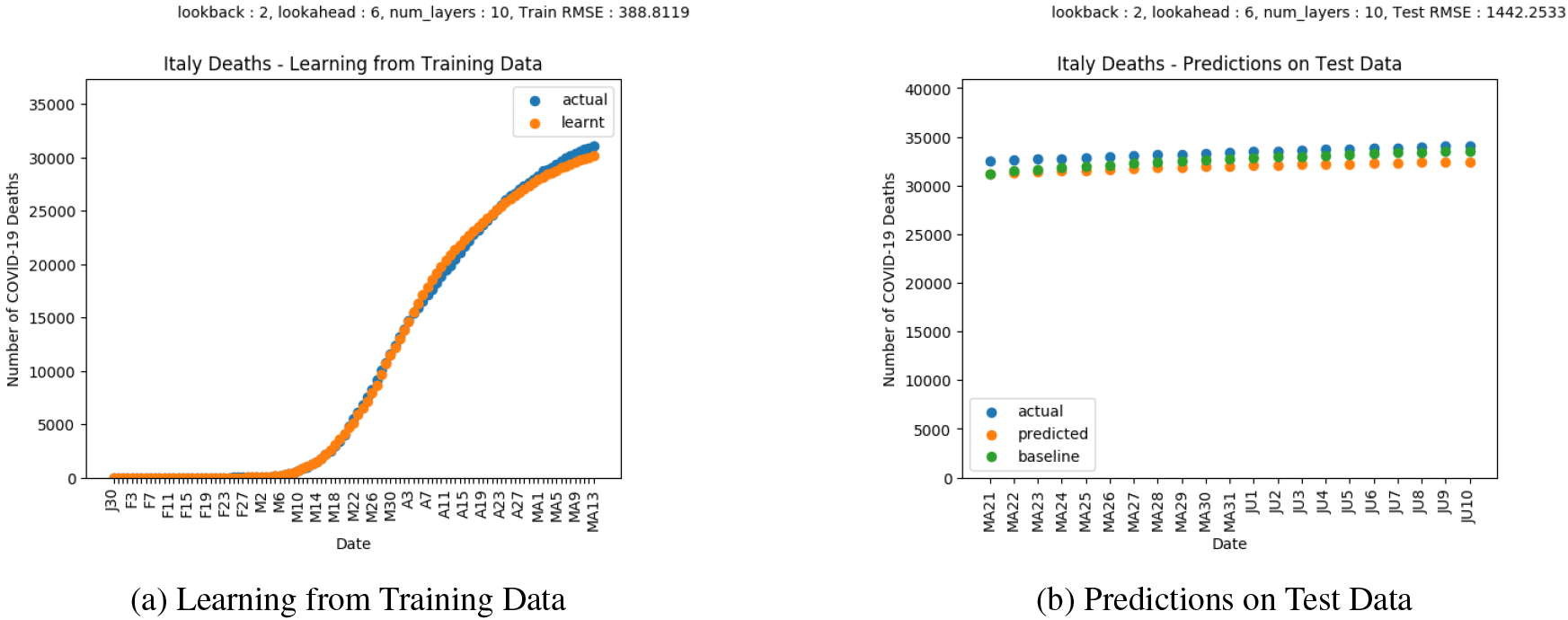
Results on COVID-19 Deaths in Italy.

Figure 6 shows the plot of predictions for COVID-19 confirmed cases in India. India enforced nationwide lockdowns in several phases following the order issued on March 24 [41]. The LSTM predictions suggest that the lockdown is yet to slow down the growing rate of COVID-19 infections as indicated by the continuing surge in COVID-19 infections.

**Figure 6:**
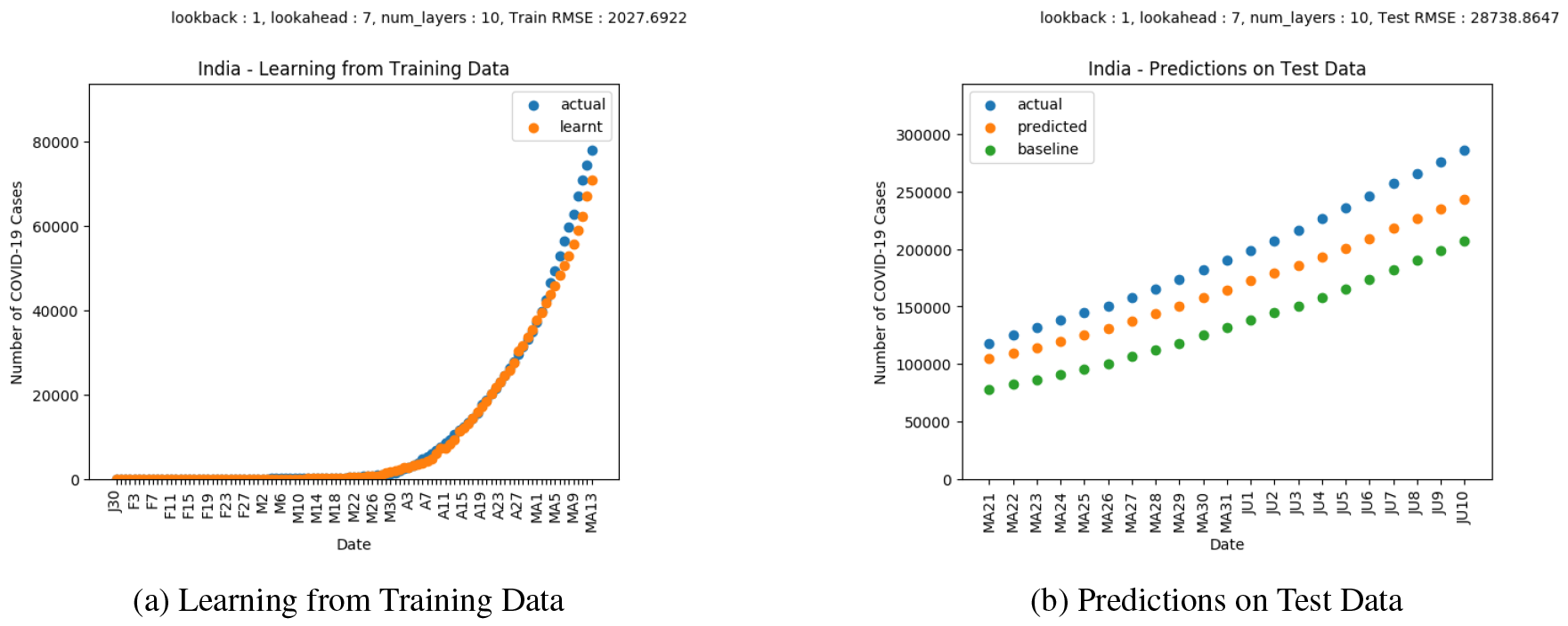
Results on COVID-19 Cases in India.

Figure 7 shows the plot of predictions for COVID-19 deaths in India. The predictions indicate that mitigation measures implemented are working to a certain extent. However, more measures may be required as the deaths showing an increasing trend.

**Figure 7:**
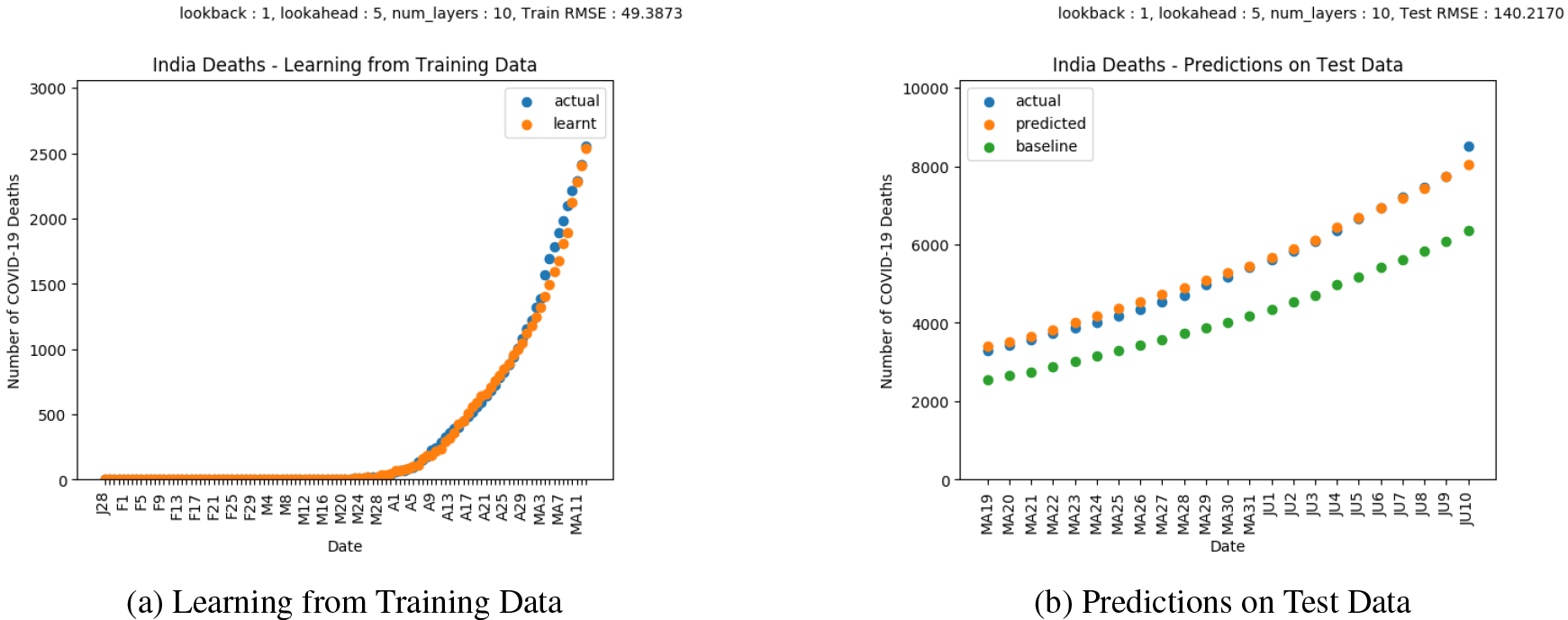
Results on COVID-19 Deaths in India.

Figure 8 shows the plot of predictions for COVID-19 confirmed cases in Japan. Japan implemented a series of measures in March 2020 to control the spread of COVID-19 [42]. The slight surge in actual cases compared to the flattening of the LSTM predictions indicates that the mitigation measures are yet to fully work.

**Figure 8:**
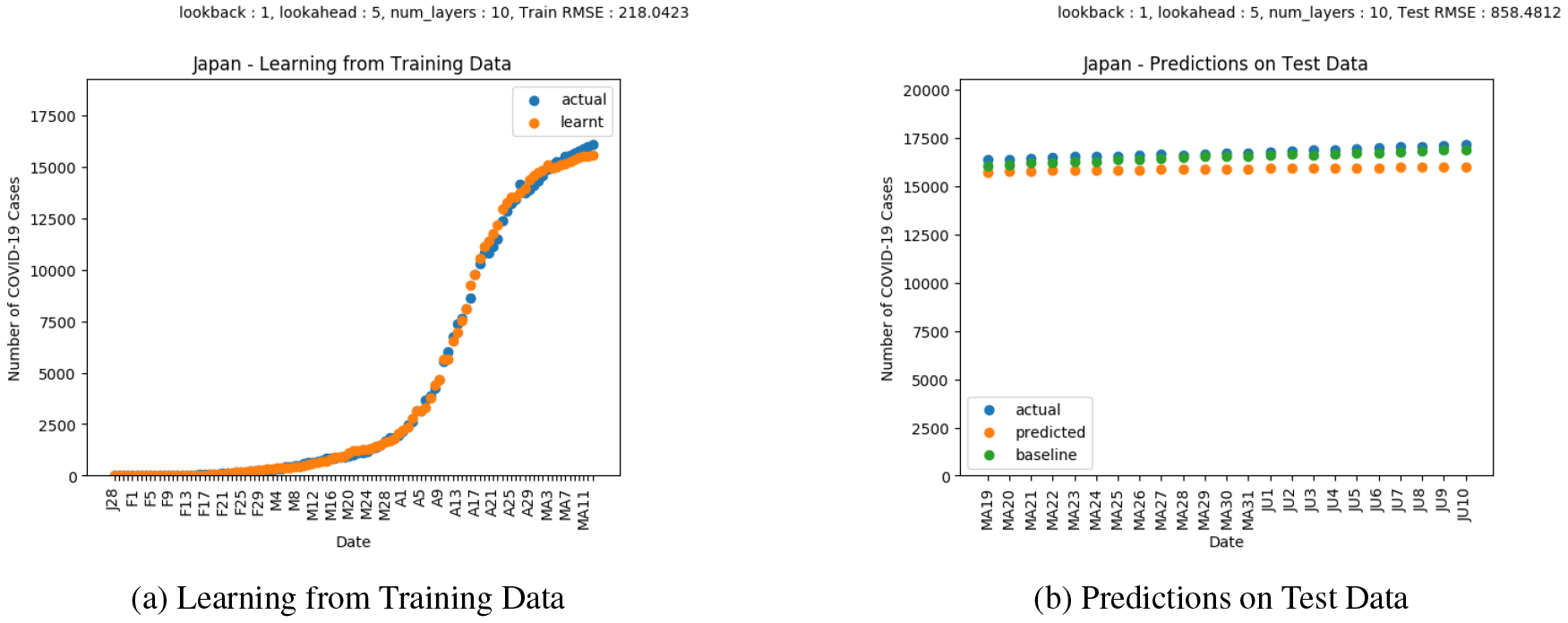
Results on COVID-19 Cases in Japan.

Figure 9 shows the plot of predictions for COVID-19 deaths in Japan. The LSTM predictions show a similar trend as the confirmed cases, however, here the COVID-19 deaths in Japan show a flattening in the curve that is also indicated by the LSTM predictions.

**Figure 9:**
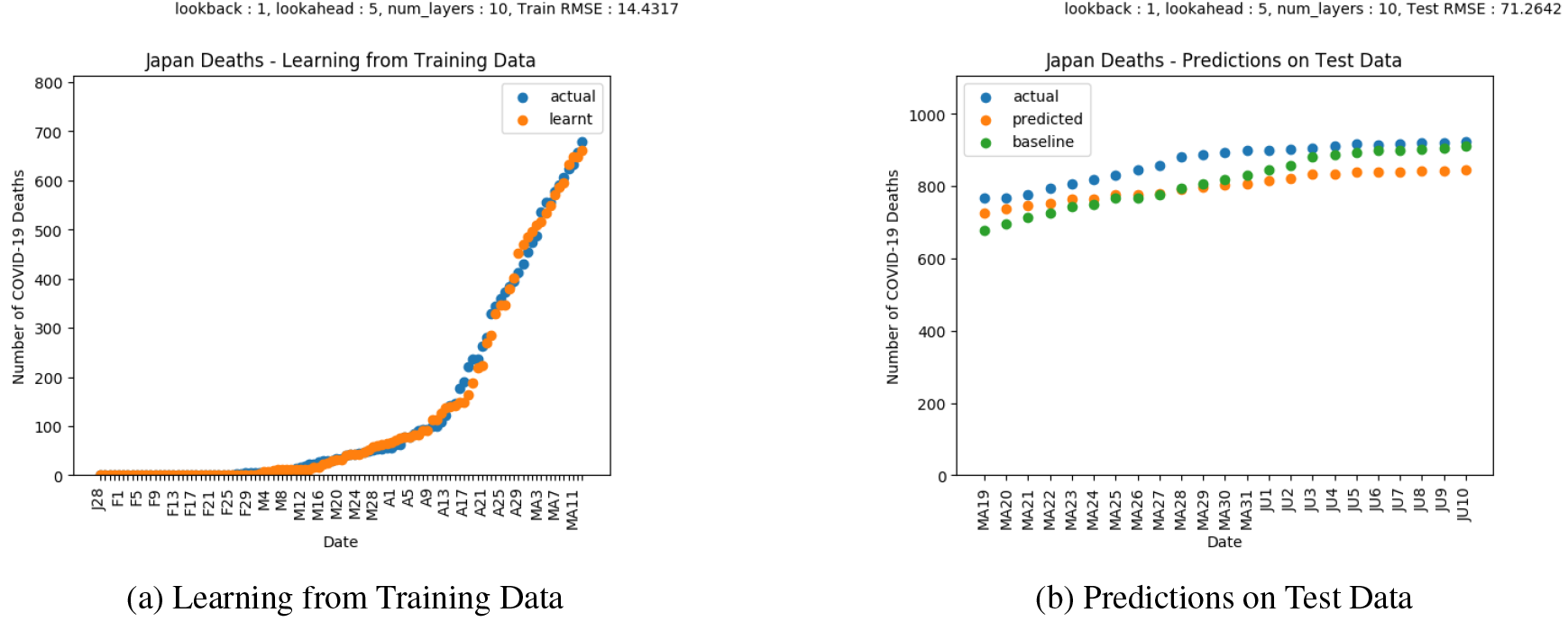
Results on COVID-19 Deaths in Japan.

#### 4.1.2 County-level Predictions

Figure 10 shows the plot of predictions for COVID-19 confirmed cases in New York City, New York, United States. New York was one of the most heavily affected areas of the United States. Counties in New York enforced lockdowns in March similar to most other places of the United States [39]. The LSTM predictions suggest that the lockdown measures have not fully worked yet as shown by the slight surge in actual cases.

**Figure 10:**
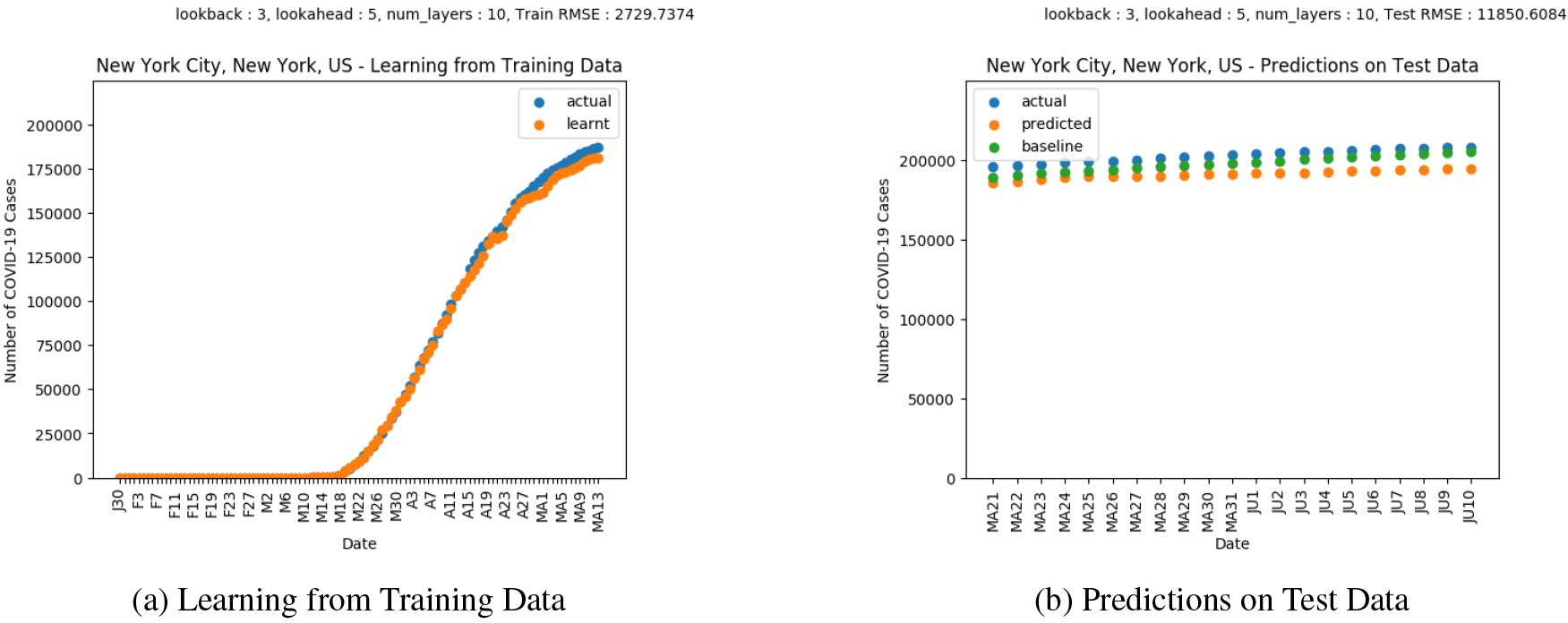
Results on COVID-19 Cases in New York City, New York, United States.

Figure 11 shows the plot of predictions for COVID-19 deaths in New York City, New York, United States. The plots show a trend similar to the infections.

**Figure 11:**
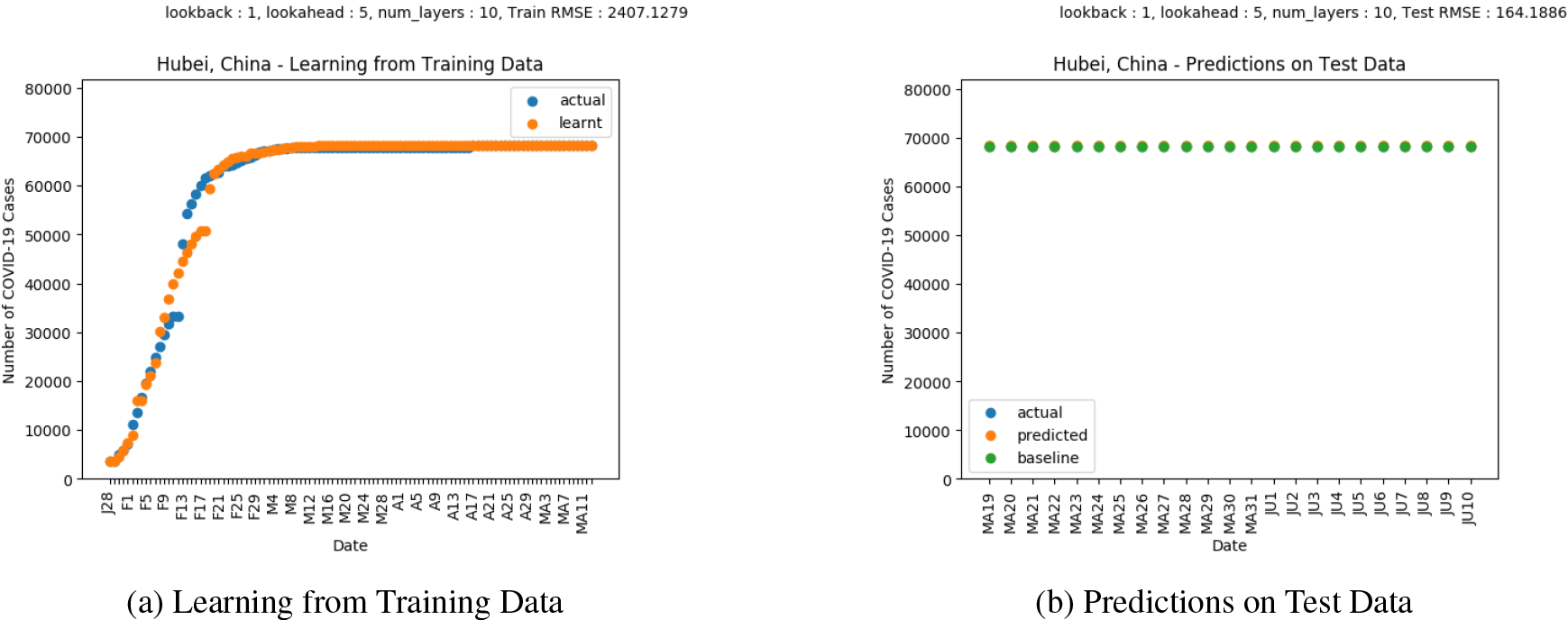
Results on COVID-19 Deaths in New York City, New York, United States.

Figure 12 shows the plot of predictions for COVID-19 confirmed cases in Hubei, China. Wuhan city in Hubei, China was the location where the first cases of COVID-19 were observed [23]. China enforced mitigation measures in January [43]. Based on both the LSTM and actual predictions, the COVID-19 cases seem to have flattened, suggesting that mitigation measures have worked.

**Figure 12:**
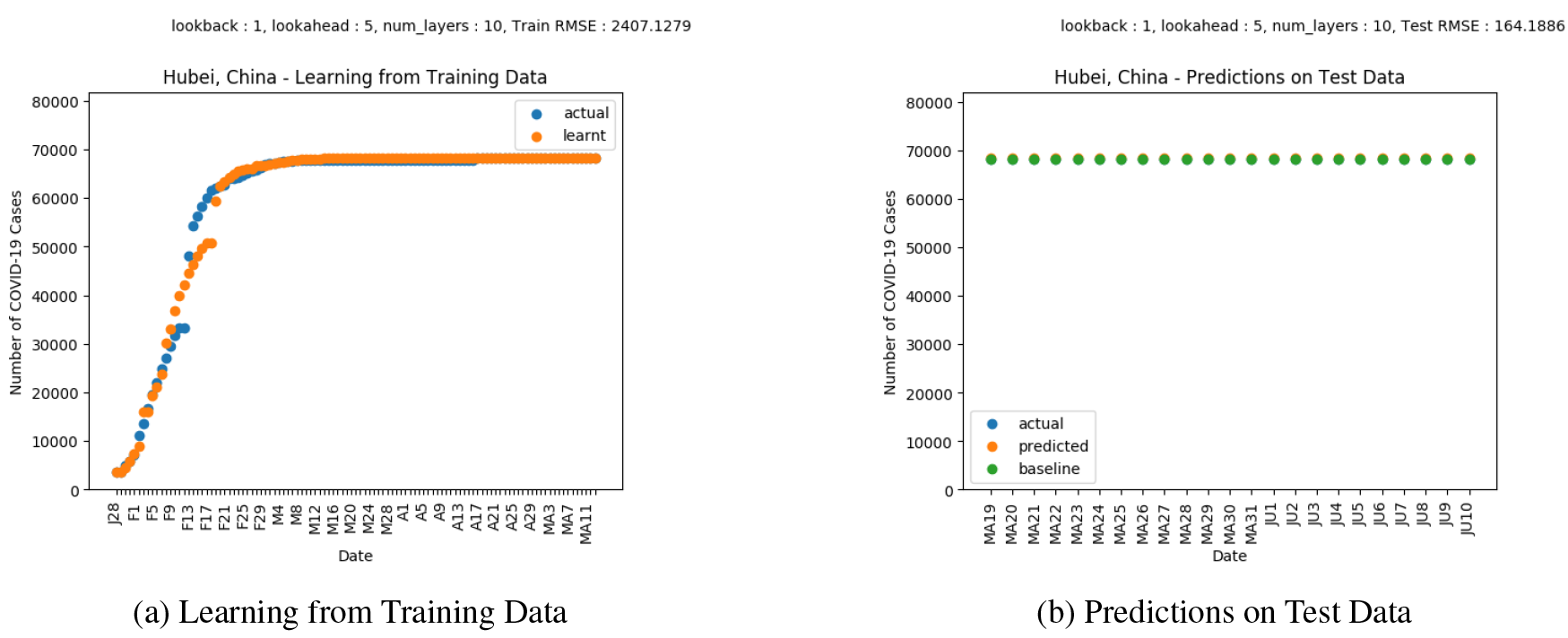
Results on COVID-19 Cases in Hubei, China.

Figure 13 shows the plot of predictions for COVID-19 deaths in Hubei, China. The trends in the LSTM predictions as well as the actual number of deaths indicate a flattening in the number of deaths. However, the difference suggests that some enforcement may still be needed before the mitigation measures are completely lifted.

**Figure 13:**
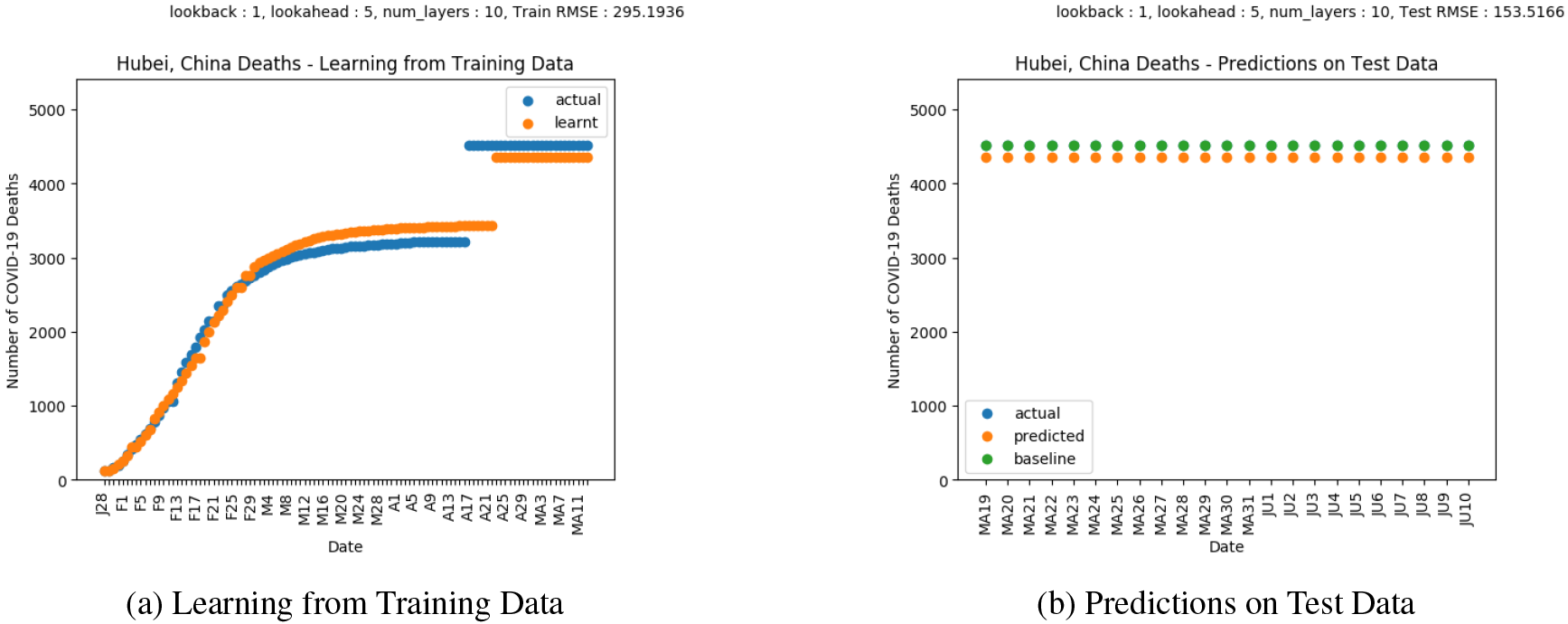
Results on COVID-19 Deaths in Hubei, China.

### 4.2 How Well are Social Distancing/Mitigation Measures Working?

We now aim at understanding how well the current mitigation measures are working. Various countries placed nationwide lockdowns, while others implemented phased strategies based on affected individual locations. The goal is to understand how social distancing had made the situation better or worse. The analyses provided are based on our proposed LSTM model. In order to do this, we consider a shorter and longer window size. In this context, we fix the *lookback* parameter and use the line of best fit on the rate of change of the difference between the smaller *lookahead* parameter of 1 and a larger *lookahead* parameter of 5. The analyses can also be carried out on individual parameters, however, for experimental purposed we choose to compute the rate on the difference in order to obtain a smoother curve for the purpose of this work. Again, the model can be used for this type of analyses for the near future as well, which will be valuable for nations or specific locations to decide whether to tighten or loosen restrictions based on the trend of how well their mitigation policies will work in the near future.

Figure 14 shows the various plots evaluating the trends of mitigation measures on rates of COVID-19 infections and deaths in five affected counties in the United States: (i) New York City, New York, (ii) Cook, Illinois, (iii) Fulton, Georgia, (iv) King, Washington, and (v) Los Angeles, California. The best_fit plots indicate the line of best fit in each figure while the LSTM_rate plots indicate the computed rate of difference of the two LSTM models as previously discussed. The line of best fit is of further interest as it provides valuable interpretations on how well the mitigation measures are working.

**Figure 14:**
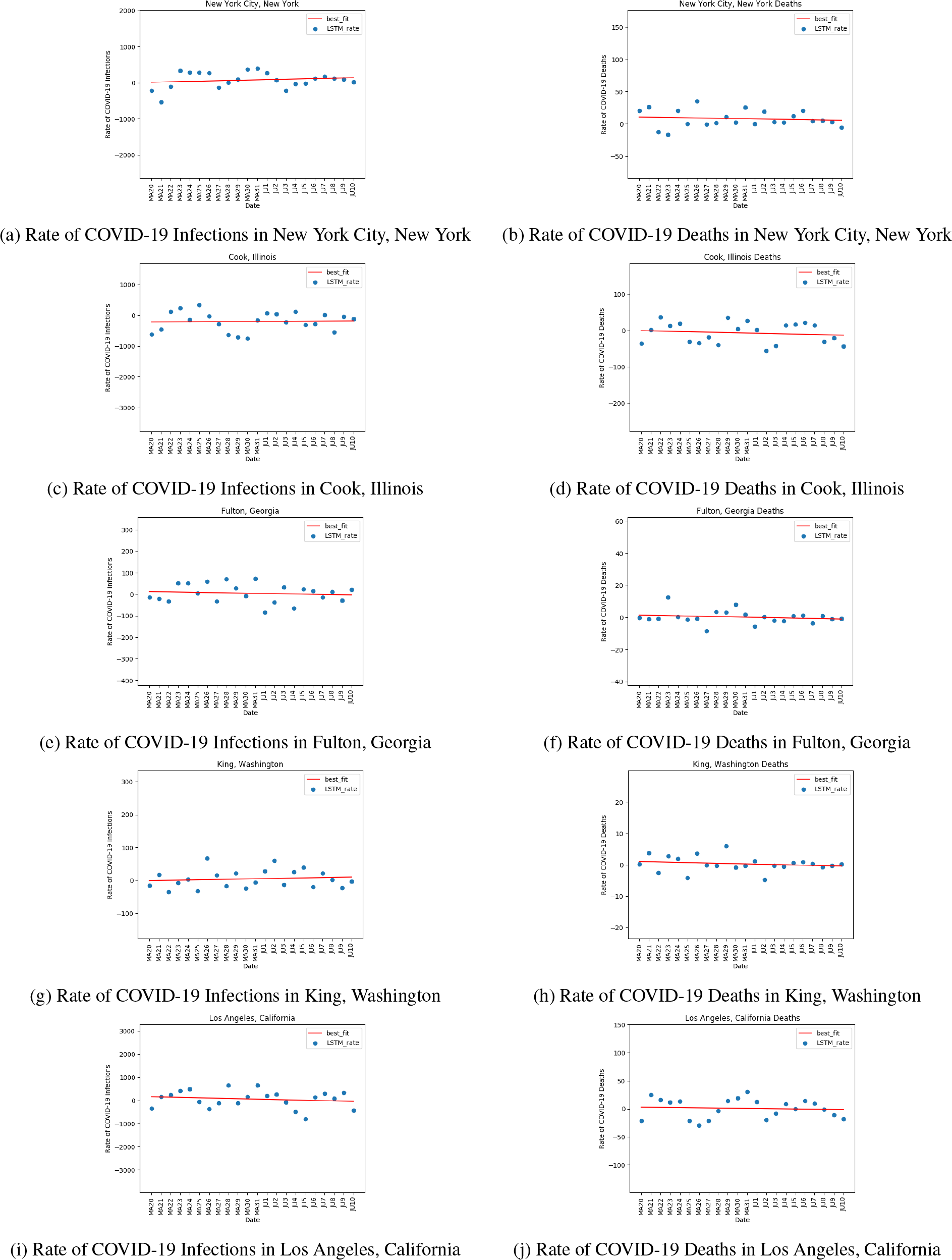
Evaluating effects of mitigation measures based on the proposed LSTM model in various counties in the United States.

In order to quantify and evaluate how well mitigation and social distancing is working, we compare the slope, intercept and RMSE values (with respect to of the line of best fit for each of the counties as shown in Table 1.

**Table 1:** Comparing effects of mitigation measures in various counties in the United States

| County | Mitigation Measures for COVID-19 Infections |  |  | Mitigation Measures for COVID-19 Deaths |  |  |
| --- | --- | --- | --- | --- | --- | --- |
|  | Slope | Intercept | RMSE | Slope | Intercept | RMSE |
| New York City, New York | 5.82 | 13.98 | 217.41 | -0.25 | 10.79 | 12.67 |
| Cook, Illinois | 1.75 | -220.14 | 306.69 | -0.58 | -0.48 | 27.97 |
| Fulton, Georgia | -0.76 | 12.57 | 41.55 | -0.12 | 1.42 | 4.06 |
| King, Washington | 0.52 | -0.64 | 27.36 | -0.07 | 1.01 | 2.33 |
| Los Angeles, California | -9.20 | 155.54 | 361.35 | -0.19 | 2.80 | 16.96 |

The slope indicates the trend of the mitigation measures – a positive value of slope indicates that the current mitigation and social distancing are not working as expected while a negative value of slope indicates that the current mitigation and social distancing are working well. In other words, a positive slope indicates an increase in the rate of infections or deaths while a negative slope indicates a decrease in the rate of infections or deaths. The intercept gives an approximation of the increase/decrease in the rate of infections or deaths depending on a positive/negative value. The RMSE gives an estimate of the stabilization of the rate of infections/deaths.

Mitigation measures New York City, New York have worked well in terms of lowering the rate of deaths but the increase in rate of infections suggests that more measures may be needed. Mitigation measures in Cook, Illinois are working in terms of the decreasing trend in deaths even though infections show an increasing trend, so prolonged mitigation measures may be needed. Mitigation measures have worked in Fulton, Georgia based on the decreasing trend of infections and deaths. However, mitigation and social distancing measures are yet to work in King, Washington as indicated by the slight increasing rate of infections even though the decreasing trend of deaths suggests that mitigation measures have worked. Mitigation measures have worked well in Los Angeles, California based on the declining rates of both infections and deaths. High fluctuations or variations in cases or deaths indicate that careful reopening measures may need to be taken as the infection rates or deaths rates have still not stabilized well.

Table 2 evaluates the success of mitigation measures based on COVID-19 infections and deaths respectively, where ✓indicates mitigation measures being successful and ✗ indicates mitigation measures not being successful. Table 2 provides an overview of the success of mitigation measures based on the analyses in Table 1. An important trend to be observed is that mitigation measures have worked based on COVID-19 deaths in all five counties in the United States that we have studied.

**Table 2:**
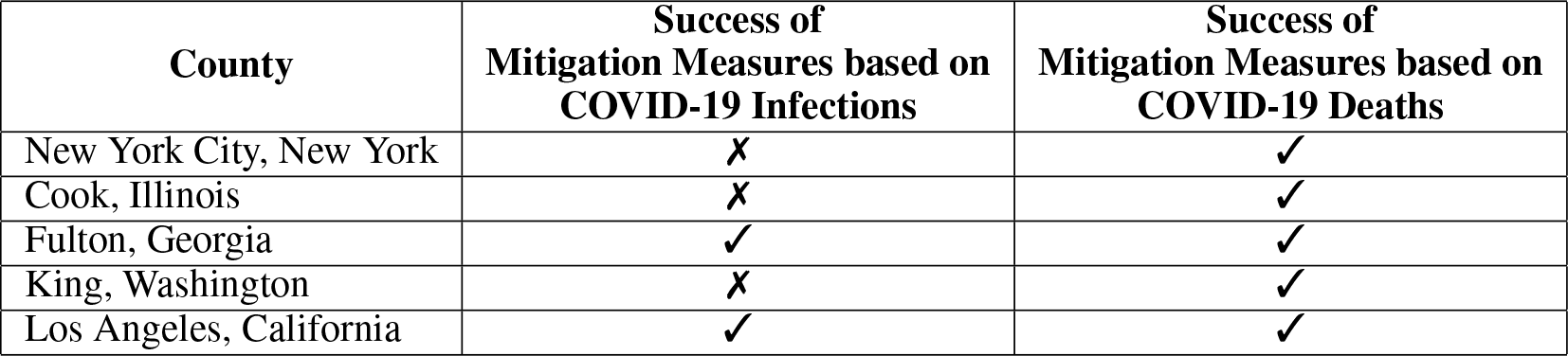
Evaluation of mitigation measures in various counties in the United States

| County | Success of Mitigation Measures based on COVID-19 Infections | Success of Mitigation Measures based on COVID-19 Deaths |
| --- | --- | --- |
| New York City, New York | ✗ | ✓ |
| Cook, Illinois | ✗ | ✓ |
| Fulton, Georgia | ✓ | ✓ |
| King, Washington | ✗ | ✓ |
| Los Angeles, California | ✓ | ✓ |

Therefore, various effects of mitigation measures have been observed in different parts of the United States which may require additional measures like rapid testing, contact tracing and phased reopening depending on the situation in various places before the entire nation resumes a normal lifestyle.

## 5 Conclusion and Future Work

The COVID-19 pandemic affected countries globally and led to an unprecedented number of infections and deaths. Countries took vital steps in mitigating the pandemic by enforcing lockdowns, social distancing, and a variety of other mitigation measures. In this paper, we propose an LSTM based learning model that can learn from the cumulative rise in COVID-19 confirmed cases and deaths and provide valuable insights on how well mitigation measures are working quantitatively in terms of the rate of infections and deaths. The predictions of the model can be helpful for countries deciding to make important decisions regarding the effects of currently implemented mitigation measures and aid in making plans for reopening various places. We provide analyses at both the country and county levels.

Future extensions of this work include implementing and understanding how this model can be transferred to studying the disease dynamics of other similar pandemics.

## Data Availability

COVID-19 Public Data: https://github.com/CSSEGISandData/COVID-19, Proposed Model Source Code: https://github.com/sayantanibasu/covid19-models

https://github.com/sayantanibasu/covid19-models

## A Code

Code for the proposed method is available here : https://github.com/sayantanibasu/covid19-models

